# Incidence, clinical characteristics and prognostic factor of patients with COVID-19: a systematic review and meta-analysis

**DOI:** 10.1101/2020.03.17.20037572

**Authors:** Chaoqun Ma, Jiawei Gu, Pan Hou, Liang Zhang, Yuan Bai, Zhifu Guo, Hong Wu, Bili Zhang, Pan Li, Xianxian Zhao

## Abstract

**Background:** Recently, Coronavirus Disease 2019 (COVID-19) outbreak started in Wuhan, China. Although the clinical features of COVID-19 have been reported previously, data regarding the risk factors associated with the clinical outcomes are lacking.

**Objectives:** To summary and analyze the clinical characteristics and identify the predictors of disease severity and mortality.

**Methods:** The PubMed, Web of Science Core Collection, Embase, Cochrane and MedRxiv databases were searched through February 25, 2020. Meta-analysis of Observational Studies in Epidemiology (MOOSE) recommendations were followed. We extracted and pooled data using random-e□ects meta-analysis to summary the clinical feature of the confirmed COVID-19 patients, and further identify risk factors for disease severity and death. Heterogeneity was evaluated using the I^2^ method and explained with subgroup analysis and meta-regression.

**Results:** A total of 30 studies including 53000 patients with COVID-19 were included in this study, the mean age was 49.8 years (95% CI, 47.5-52.2 yrs) and 55.5% were male. The pooled incidence of severity and mortality were 20.2% (95% CI, 15.1-25.2%) and 3.1% (95% CI, 1.9-4.2%), respectively. The predictor for disease severity included old age (≥ 50 yrs, odds ratio [OR] = 2.61; 95% CI, 2.29-2.98), male (OR =1.348, 95% CI, 1.195-1.521), smoking (OR =1.734, 95% CI, 1.146-2.626) and any comorbidity (OR = 2.635, 95% CI, 2.098-3.309), especially chronic kidney disease (CKD, OR = 6.017; 95% CI, 2.192-16.514), chronic obstructive pulmonary disease (COPD, OR = 5.323; 95% CI, 2.613-10.847) and cerebrovascular disease (OR = 3.219; 95% CI, 1.486-6.972). In terms of laboratory results, increased lactate dehydrogenase (LDH), C-reactive protein (CRP) and D-dimer and decreased blood platelet and lymphocytes count were highly associated with severe COVID-19 (all for P < 0.001). Meanwhile, old age (≥ 60 yrs, RR = 9.45; 95% CI, 8.09-11.04), followed by cardiovascular disease (RR = 6.75; 95% CI, 5.40-8.43) hypertension (RR = 4.48; 95% CI, 3.69-5.45) and diabetes (RR = 4.43; 95% CI, 3.49-5.61) were found to be independent prognostic factors for the COVID-19 related death.

**Conclusions:** To our knowledge, this is the first evidence-based medicine research to explore the risk factors of prognosis in patients with COVID-19, which is helpful to identify early-stage patients with poor prognosis and adapt effective treatment.

## Introduction

Coronaviruses are a family of viruses that widely exist in nature and can infect both humans and animals^1^. At present, there are six recognized types of coronavirus that can cause infection in humans, leading to pneumonia, injury in the digestive tracts, kidney failure and even death. Two of the coronavirus family have been reported to cause deadly infections including Middle East Respiratory Syndrome (MERS) and Severe Acute Respiratory Syndrome (SARS)^2,3^. Recently, pneumonia caused by a novel coronavirus (SARS-CoV-2), also termed as Coronavirus Disease 2019 (COVID-19) was first reported in China’s Wuhan City, Hubei Province in December 2019 and the disease spread rapidly in China and even around the world^4^.

Compared with two other types of coronaviruses, the present new coronavirus is spreading far more quickly and has higher contagiousness^5^. As of March 17, 2020, a total of 187, 361 COVID-19 cases in 151 countries have been confirmed, which almost 22.2 times the number of people infected by the SARS in 2003. Although COVID-19 has a relatively low mortality rate, it can be highly deadly and lethal, especially in high-risk patients^6^. The reported incidence of COVID-19 accompanied with underlying comorbidities in the literature were up to 26.0%, and most of them (65.3%) had cardiovascular and cerebrovascular diseases. Patients infected with SARS-CoV-2 who already have underlying diseases were at increased the risk of severe illnesses and death^7^.

And more worrying, there is no vaccine and specific treatment available for this novel coronavirus as of now^8^. Therefore, it is necessary to identify potential risk factors for predicting the disease progression and severity for COVID-19. Meanwhile, early monitoring the predictive indicators may also increase the efficiency of treatment and improve the prognosis of COVID-19. Thus, the aim of our study is to perform a systematic review and meta-analysis of clinical characteristics to explore the risk factors of COVID-19-associated severe illness and death, and first time to compare the differences of those predictors between COVID-19, SARS and MERS.

## Methods

Our study was performed according to the Preferred Reporting Items for Systematic Reviews and Meta-Analyses (PRISMA) statement and Meta-analysis of Observational Studies in Epidemiology (MOOSE) reporting guidelines^9,10^. In eMethods in the Supplement, we describe the detailed definitions of COVID-19 laboratory confirmed cases and severe illness.

### Search strategy and study selection

PubMed, Embase, Cochrane, the Web of Science Core Collection (Clarivate Analytics), and MedRxiv databases were used for searching articles published until February 25, 2020 using the following keywords: “coronavirus”, “nCoV”, “HCoV”, “Wuhan”, “2019” “SARS-CoV-2”, “COVID*”, “NCP*”, “China”, “clinical”, “outcome”, “sever*”, “death”, “fatali*” and “mortalit*” alone and in combination. Detailed search strategies were presented in eMethods in the Supplement. After removing duplicates, three reviewers (C.M. & J.G. & P. H) were assigned to independently screen the titles and abstracts and then examine the full text, and any questions with conflict were resolved by the senior authors (P. L. and X. Z.). Inclusion criteria were as follows: (1) any study that gives information about the clinical characteristics or demographic or outcome of the infectious disease, (2) restriction language to English only, and (3) studies that allowed us to stratify the risk of severe or fatal COVID-19 by demographic or medical condition were preferred. Exclusion criteria were (1) data that could not be reliably extracted, (2) editorials, comments, expert opinions, case reports or articles with number of patients ≤ 10, (3) studies with special populations (eg, only focused on family clusters or severe or death cases).

### Data extraction and quality assessment

Two authors (C.M. and J.G.) independently extracted the data using a predesigned spreadsheet, including author’s name, publication year, study period, geographical region, study design, epidemiological information, sample size, baseline and clinical characteristics, laboratory results, severity and outcomes. For the study cohort divided according to disease severity, data of all patients, severe and non-severe patients were collected, respectively. For further meta-analysis, categorical variables like sex, comorbidities, symptoms or endpoint events, number were dealt with as dichotomous variables, while for continuous variables such as age, laboratory results or timelines of illness, different types of measurement including median (range) or median (IQR, interquartile range) were transferred to the form of mean (SD, standard deviation)^10^. Along with data extraction, study quality was assessed using the Quality Assessment Forms recommended by Agency for Healthcare Research and Quality (AHRQ) for cross-sectional study (eMethods in supplement)^11^. Conflicts on the assessments were resolved either by consensus or by the adjudicator (P.L. and X. Z.).

### Data analysis

Firstly, to obtain summary e□ect estimate for each clinical variable, including case severity rate (CSR), case fatality rate (CFR), male proportion, mean age, pooled value of lymphocyte count and timeline of COVID-19 confirmed patients, random e□ects meta-analysis was used because high variability between studies was expected. Heterogeneity was evaluated using the I^2^ statistic. Cut-off values of 25%, 50%, and 75% indicated low, moderate, and high heterogeneity, respectively^12^. We visualized the results with forest plots. Secondly, to identify the risk factors for severity, pooled odds ratios (ORs) with 95% CIs were estimated with the dichotomous method, and mean difference (MD) with 95% CIs between severe and non-severe cases were calculated with continuous method, Fixed effects model was used when I^2^ < 50%, and random effects models otherwise. Regarding the COVID-19 related death, we conducted logistic regression model to calculate relative risks (RR) with 95% CIs using the data of Chinese Center for Disease Control, which included 44672 laboratory confirmed patients. Thirdly, to explore potential sources of heterogeneity, we conducted subgroup analysis and random-e□ects meta-regression. Variables significant in univariable meta-regression (*P* < 0.05) were included in multivariable meta-regression. Next, sensitivity analyses were performed by systematically removing each study in turn to explore its effect on outcome^13^.

Finally, to investigate the risk of publication bias, we applied the Begger test, Egger test and the test used by Peters et al and visually inspected the funnel plots^14^. All analyses were performed using Review Manager (version5.3), Stata (version 15) and R (version 3.5.3), RStudio (version 1.2.1335) and Comprehensive Meta-analysis (version 3.3).

## Results

### Literature Search and Study Characteristics

We identified 854 studies, of which 30 were eligible for our analysis including 53,000 COVID-19 confirmed patients (Figure *1*)^1,7,15-38^. All of them were retrospective, observational studies (19 single-center and 11 multi-center studies), which were performed between December 2019 and February 19, 2020. The majority of studies were conducted in Wuhan (13, 43.3%) and other cities in China (11, 36.7%), 2 from nationwide and 3 from other countries including United States, Australia and Korea. To avoid any overlap of cases, the nationwide study of Chinese Centers for Disease Control (CDC) including 44672 confirmed cases was only used for identifying COVID-19 related death risk factors. The male to female sex ratio was 1.25, with an overall average age of 49.8 years (95% CI, 47.5-52.2). Five studies included the information of COVID-19 infected medical staff (Table *1*).

**Table 1.**
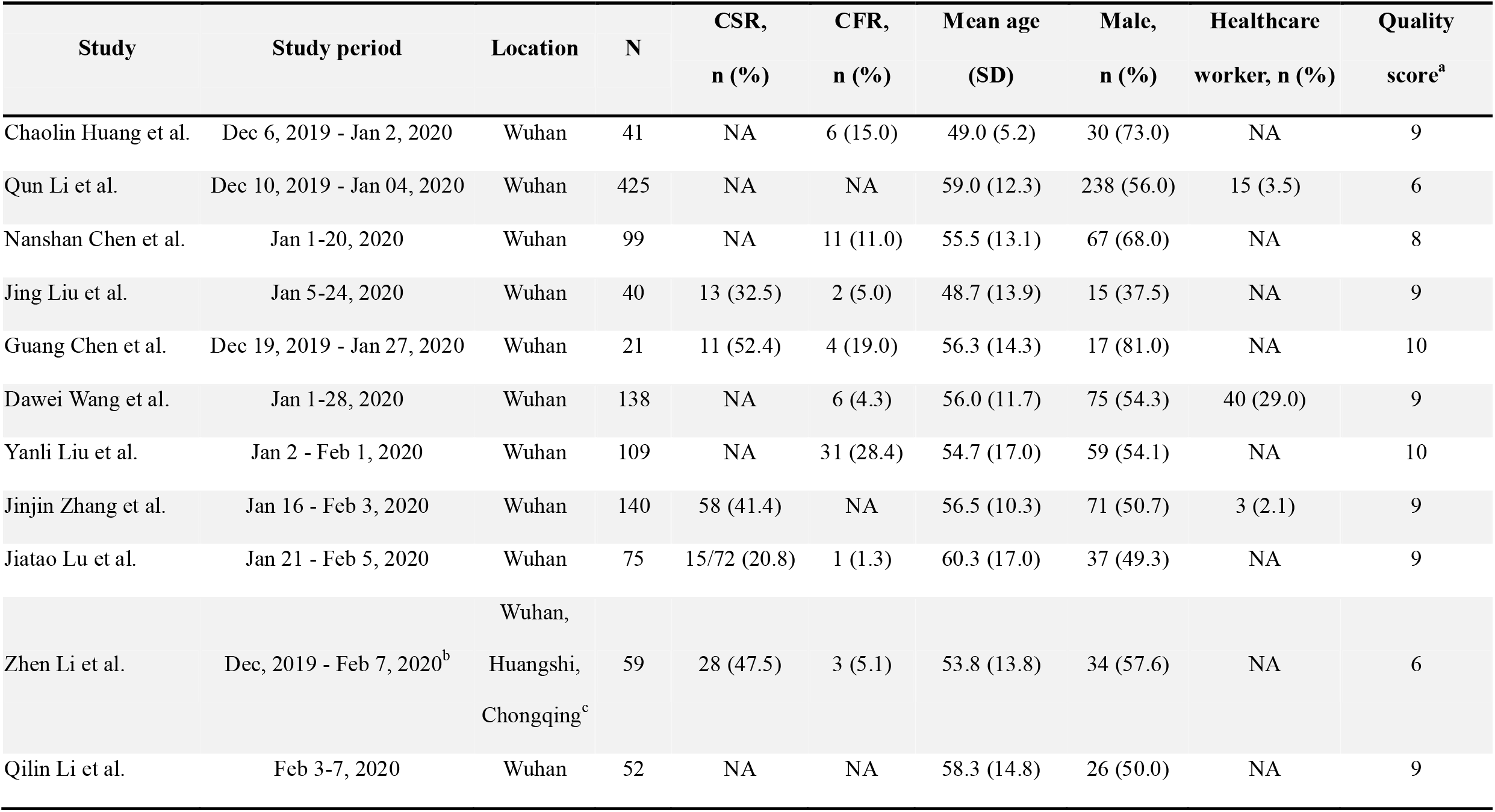
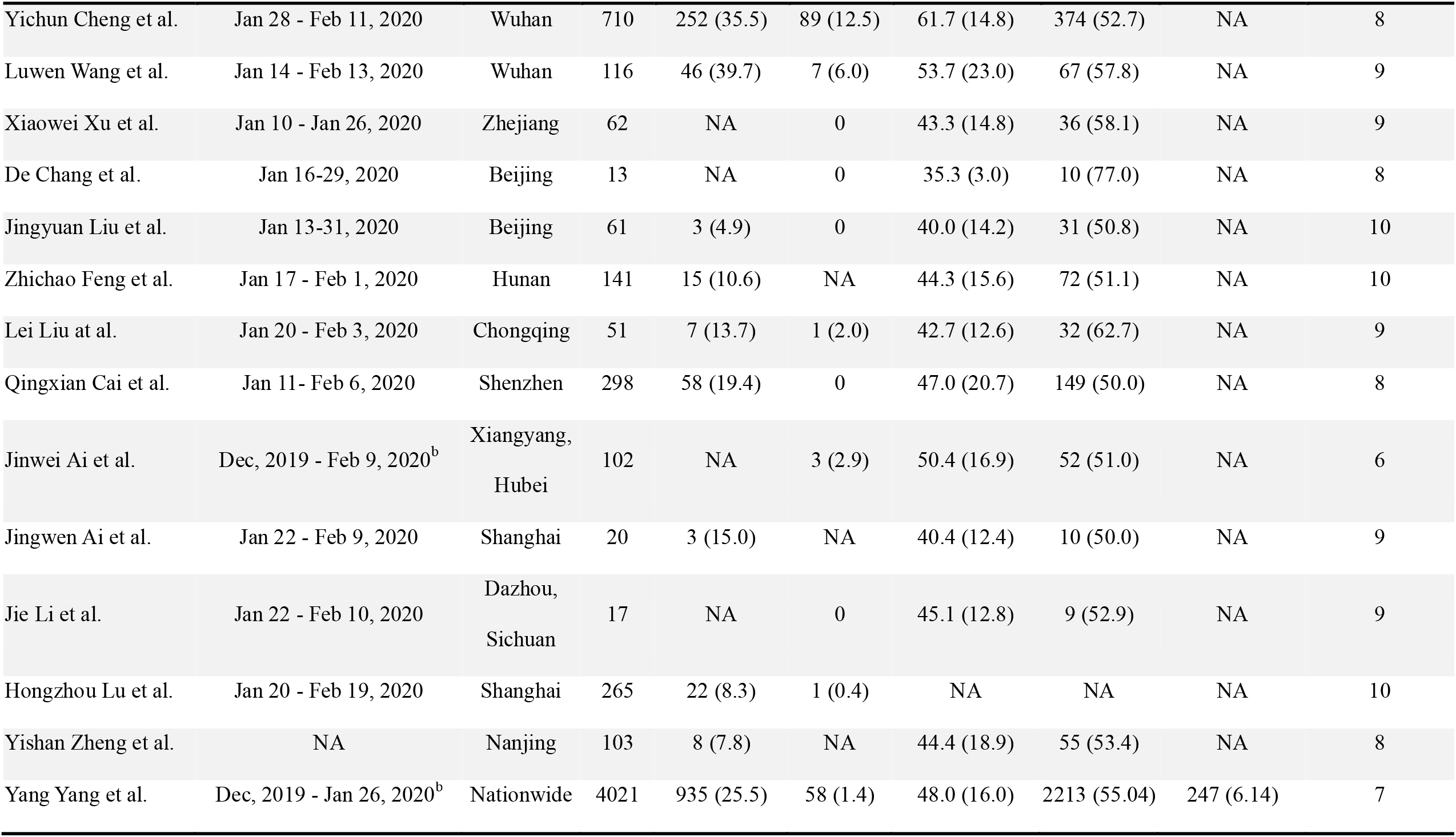
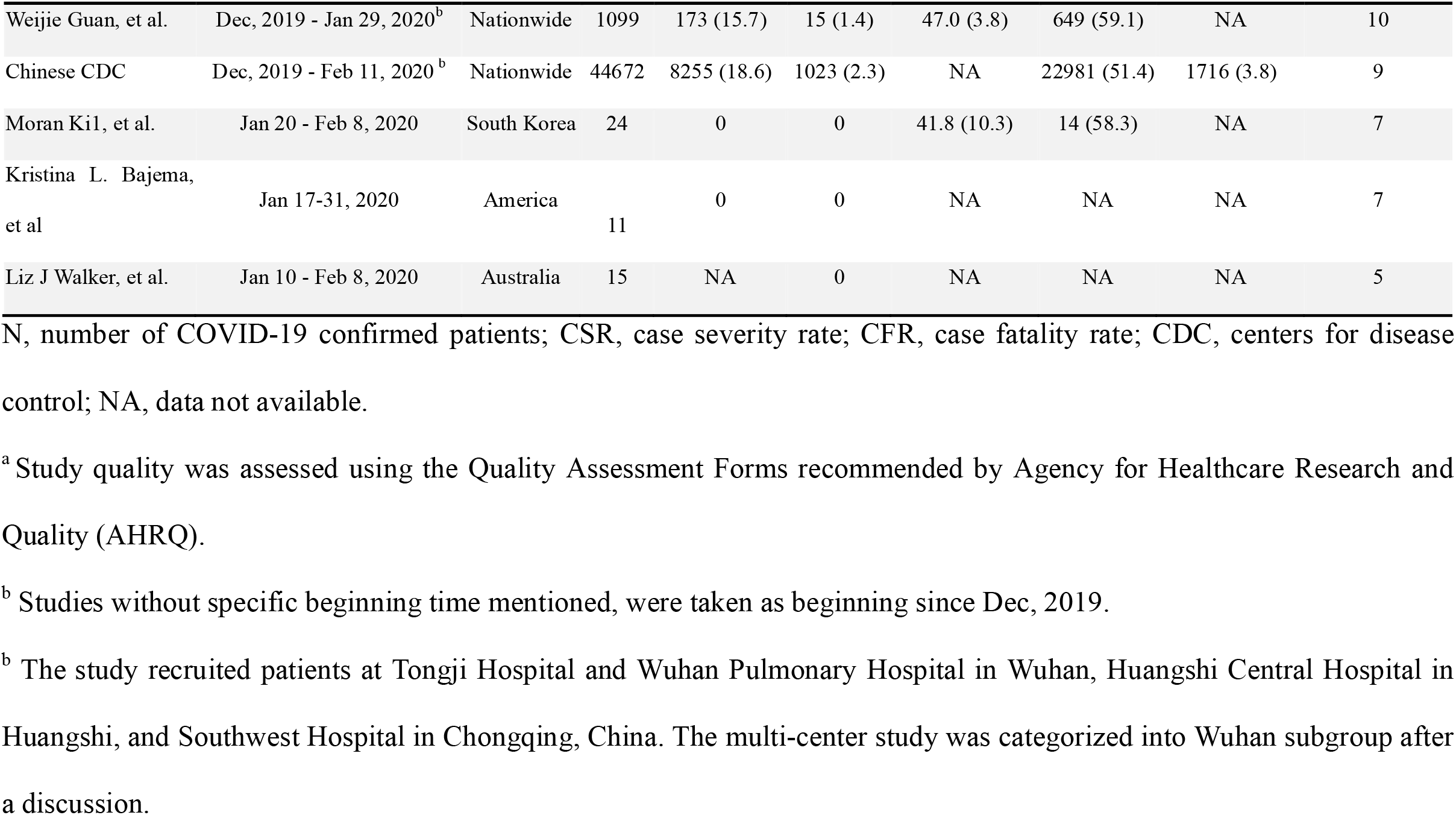
Study Characteristics

| Study | Study period | Location | N | CSR,<br>n (%) | CFR,<br>n (%) | Mean age<br>(SD) | Male,<br>n (%) | Healthcare<br>worker, n (%) | Quality<br>score<sup>a</sup> |
| --- | --- | --- | --- | --- | --- | --- | --- | --- | --- |
| Chaolin Huang et al. | Dec 6, 2019 - Jan 2, 2020 | Wuhan | 41 | NA | 6 (15.0) | 49.0 (5.2) | 30 (73.0) | NA | 9 |
| Qun Li et al. | Dec 10, 2019 - Jan 04, 2020 | Wuhan | 425 | NA | NA | 59.0 (12.3) | 238 (56.0) | 15 (3.5) | 6 |
| Nanshan Chen et al. | Jan 1-20, 2020 | Wuhan | 99 | NA | 11 (11.0) | 55.5 (13.1) | 67 (68.0) | NA | 8 |
| Jing Liu et al. | Jan 5-24, 2020 | Wuhan | 40 | 13 (32.5) | 2 (5.0) | 48.7 (13.9) | 15 (37.5) | NA | 9 |
| Guang Chen et al. | Dec 19, 2019 - Jan 27, 2020 | Wuhan | 21 | 11 (52.4) | 4 (19.0) | 56.3 (14.3) | 17 (81.0) | NA | 10 |
| Dawei Wang et al. | Jan 1-28, 2020 | Wuhan | 138 | NA | 6 (4.3) | 56.0 (11.7) | 75 (54.3) | 40 (29.0) | 9 |
| Yanli Liu et al. | Jan 2 - Feb 1, 2020 | Wuhan | 109 | NA | 31 (28.4) | 54.7 (17.0) | 59 (54.1) | NA | 10 |
| Jinjin Zhang et al. | Jan 16 - Feb 3, 2020 | Wuhan | 140 | 58 (41.4) | NA | 56.5 (10.3) | 71 (50.7) | 3 (2.1) | 9 |
| Jiatao Lu et al. | Jan 21 - Feb 5, 2020 | Wuhan | 75 | 15/72 (20.8) | 1 (1.3) | 60.3 (17.0) | 37 (49.3) | NA | 9 |
| Zhen Li et al. | Dec, 2019 - Feb 7, 2020<sup>b</sup> | Wuhan,<br>Huangshi,<br>Chongqing<sup>c</sup> | 59 | 28 (47.5) | 3 (5.1) | 53.8 (13.8) | 34 (57.6) | NA | 6 |
| Qilin Li et al. | Feb 3-7, 2020 | Wuhan | 52 | NA | NA | 58.3 (14.8) | 26 (50.0) | NA | 9 |
| Yichun Cheng et al. | Jan 28 - Feb 11, 2020 | Wuhan | 710 | 252 (35.5) | 89 (12.5) | 61.7 (14.8) | 374 (52.7) | NA | 8 |
| Luwen Wang et al. | Jan 14 - Feb 13, 2020 | Wuhan | 116 | 46 (39.7) | 7 (6.0) | 53.7 (23.0) | 67 (57.8) | NA | 9 |
| Xiaowei Xu et al. | Jan 10 - Jan 26, 2020 | Zhejiang | 62 | NA | 0 | 43.3 (14.8) | 36 (58.1) | NA | 9 |
| De Chang et al. | Jan 16-29, 2020 | Beijing | 13 | NA | 0 | 35.3 (3.0) | 10 (77.0) | NA | 8 |
| Jingyuan Liu et al. | Jan 13-31, 2020 | Beijing | 61 | 3 (4.9) | 0 | 40.0 (14.2) | 31 (50.8) | NA | 10 |
| Zhichao Feng et al. | Jan 17 - Feb 1, 2020 | Hunan | 141 | 15 (10.6) | NA | 44.3 (15.6) | 72 (51.1) | NA | 10 |
| Lei Liu at al. | Jan 20 - Feb 3, 2020 | Chongqing | 51 | 7 (13.7) | 1 (2.0) | 42.7 (12.6) | 32 (62.7) | NA | 9 |
| Qingxian Cai et al. | Jan 11- Feb 6, 2020 | Shenzhen | 298 | 58 (19.4) | 0 | 47.0 (20.7) | 149 (50.0) | NA | 8 |
| Jinwei Ai et al. | Dec, 2019 - Feb 9, 2020<sup>b</sup> | Xiangyang,<br>Hubei | 102 | NA | 3 (2.9) | 50.4 (16.9) | 52 (51.0) | NA | 6 |
| Jingwen Ai et al. | Jan 22 - Feb 9, 2020 | Shanghai | 20 | 3 (15.0) | NA | 40.4 (12.4) | 10 (50.0) | NA | 9 |
| Jie Li et al. | Jan 22 - Feb 10, 2020 | Dazhou,<br>Sichuan | 17 | NA | 0 | 45.1 (12.8) | 9 (52.9) | NA | 9 |
| Hongzhou Lu et al. | Jan 20 - Feb 19, 2020 | Shanghai | 265 | 22 (8.3) | 1 (0.4) | NA | NA | NA | 10 |
| Yishan Zheng et al. | NA | Nanjing | 103 | 8 (7.8) | NA | 44.4 (18.9) | 55 (53.4) | NA | 8 |
| Yang Yang et al. | Dec, 2019 - Jan 26, 2020<sup>b</sup> | Nationwide | 4021 | 935 (25.5) | 58 (1.4) | 48.0 (16.0) | 2213 (55.04) | 247 (6.14) | 7 |
| Weijie Guan, et al. | Dec, 2019 - Jan 29, 2020<sup>b</sup> | Nationwide | 1099 | 173 (15.7) | 15 (1.4) | 47.0 (3.8) | 649 (59.1) | NA | 10 |
| Chinese CDC | Dec, 2019 - Feb 11, 2020<sup>b</sup> | Nationwide | 44672 | 8255 (18.6) | 1023 (2.3) | NA | 22981 (51.4) | 1716 (3.8) | 9 |
| Moran Ki1, et al. | Jan 20 - Feb 8, 2020 | South Korea | 24 | 0 | 0 | 41.8 (10.3) | 14 (58.3) | NA | 7 |
| Kristina L. Bajema,<br>et al | Jan 17-31, 2020 | America | 11 | 0 | 0 | NA | NA | NA | 7 |
| Liz J Walker, et al. | Jan 10 - Feb 8, 2020 | Australia | 15 | NA | 0 | NA | NA | NA | 5 |

N, number of COVID-19 confirmed patients; CSR, case severity rate; CFR, case fatality rate; CDC, centers for disease control; NA, data not available.
<sup>a</sup> Study quality was assessed using the Quality Assessment Forms recommended by Agency for Healthcare Research and Quality (AHRQ).
<sup>b</sup> Studies without specific beginning time mentioned, were taken as beginning since Dec, 2019.
<sup>b</sup> The study recruited patients at Tongji Hospital and Wuhan Pulmonary Hospital in Wuhan, Huangshi Central Hospital in Huangshi, and Southwest Hospital in Chongqing, China. The multi-center study was categorized into Wuhan subgroup after a discussion.

**Figure 1.**
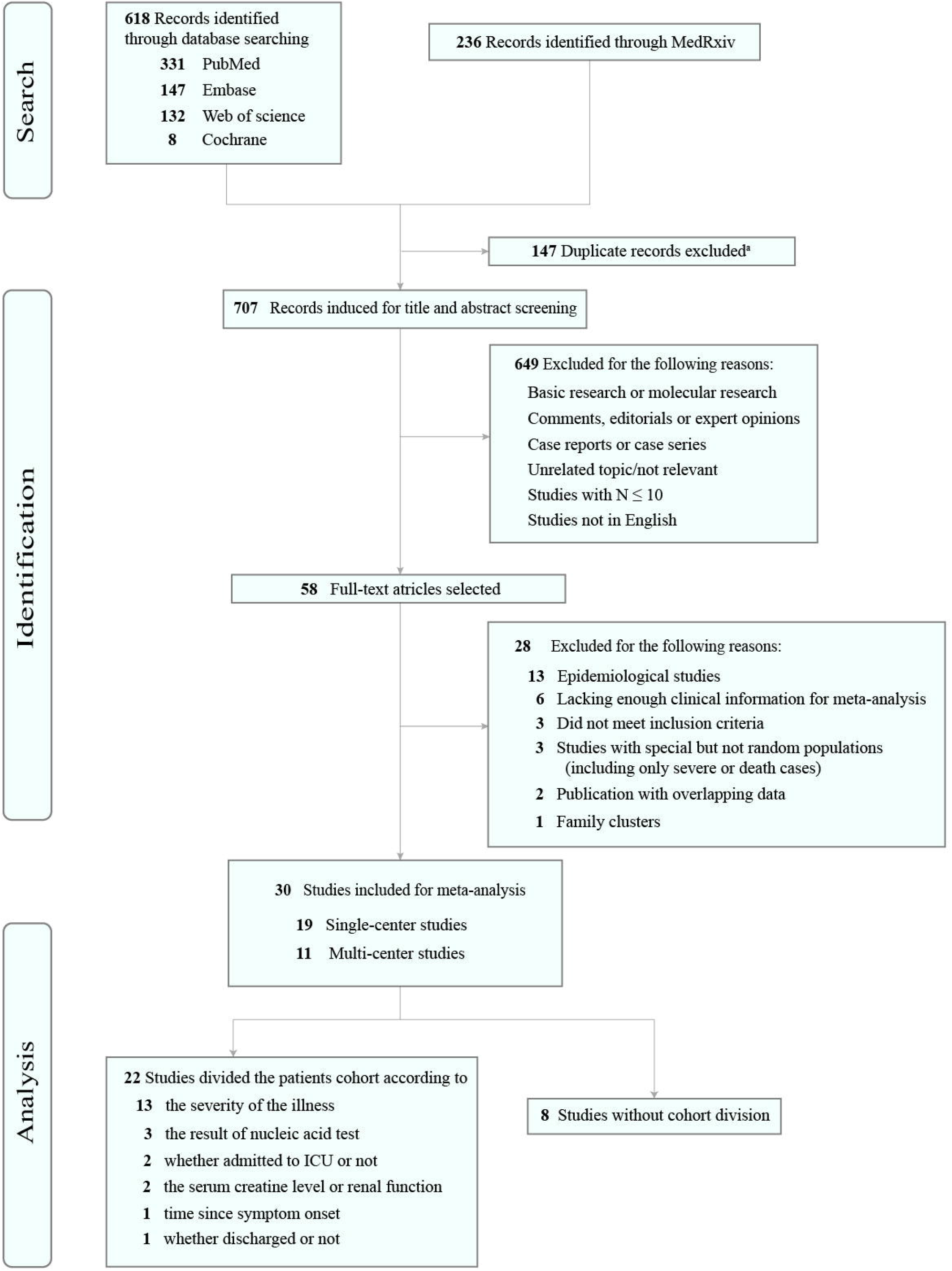
PRISMA Diagram of Study Selection. N, the number of COVID-19 confirmed patients; ICU, intensive care unit. a EndNote software (Clarivate Analytics) was used to remove duplicates.

### Case Severity Rate and Case Fatality Rate

18 studies listed the number of severe cases, with pooled CSR 20.2% (95% CI, 15.1-25.2%, n = 18, I^2^ = 92%). The proportion of severe illness in Wuhan subgroup was higher than outside of Wuhan in China (36.9%; [95% CI, 26.7-47.0%]; n = 7 vs. 10.9%; [95% CI, 6.7-15.1%]; n = 7, *P* < 0.001]). Of note, the overall case fatality rate (CFR) was 3.1% (95% CI, 1.9-4.2%, n = 23, I^2^ = 75%). Subgroup analysis showed a significantly higher CFR in Wuhan than outside Wuhan (9.5%; [95% CI, 5.2-13.8%]; n = 10 vs. 0.2%; [95% CI, 0-0.5%]; n = 8, *P* < 0.001) (Figure *1*).

Univariate meta-regression showed that compared with Wuhan, CSR and CFR in other areas were significantly lower (all for *P* < 0.0001), which was consistent with the subgroup analysis. Although not statistically significant, CSR and CFR showed a decreasing trend over time. In addition, onset-to-admission time were identified closely correlation with CSR (4.99% per increase in days, *P* = 0.0047) and CFR (1.97% per increase in days, *P* < 0.0001), suggesting shortening the onset-to-admission time favored COVID-19 related outcomes. Multivariate meta-regression confirmed the close correlation between onset-to-admission time and CFR (1.27% per increase in days, *P* = 0.0263). (Table *2* and Figure *3*).

**Table 2.** Meta-regression Analysis for CSR and CFR

|  | Univariate meta-regression |  |  | Multivariate meta-regression<sup>a</sup> |  |  |
| --- | --- | --- | --- | --- | --- | --- |
|  | Coefficient (95% CI) | z value | p value | Coefficient (95% CI) | z value | p value |
| CSR |  |  |  |  |  |  |
| Geographic region |  |  |  |  |  |  |
| Wuhan | Reference | 10.1055 | Reference | Reference | 3.3328 | Reference |
| Outside Wuhan in China | -0.2472 (-0.3417 to -0.1528) | -5.1316 | **<0.0001** | -0.1317 (-0.3150 to 0.0517) | -1.4074 | 0.1593 |
| Nationwide | -0.1516 (-0.2813 to -0.0219) | -2.2909 | **0.0220** | -0.0122 (-0.2162 to 0.2405) | 0.1044 | 0.9169 |
| Outside China | -0.3581 (-0.5004 to -0.2159) | -4.9355 | **<0.0001** |  |  |  |
| Observation duration<sup>b</sup> | -0.0048 (-0.0102 to 0.0006) | -1.7499 | 0.0801 | - | - | - |
| Onset-to-admission time<sup>c</sup> | 0.0499 (0.0153 to 0.0845) | 2.8289 | **0.0047** | 0.0275 (-0.0189 to 0.0739) | 1.1606 | 0.2458 |
| CFR |  |  |  |  |  |  |
| Geographic region |  |  |  |  |  |  |
| Wuhan | Reference | 8.5225 | Reference | Reference | 1.0729 | Reference |
| Outside Wuhan in China | -0.0712 (-0.0935 to -0.0488) | -6.2345 | **<0.0001** | -0.0450 (-0.0922 to 0.0022) | -1.8686 | 0.0617 |
| Nationwide | -0.0627 (-0.0894 to -0.0359) | -4.5955 | **<0.0001** | -0.0381 (-0.0949 to 0.0187) | -1.3156 | 0.1883 |
| Outside China | -0.0767 (-0.1264 to -0.0269) | -3.0214 | **0.0025** |  |  |  |
| Observation duration<sup>b</sup> | -0.0002 (-0.0013 to 0.0009) | -0.3710 | 0.7106 | - | - | - |
| Onset-to-admission time<sup>c</sup> | 0.0197 (0.0133 to 0.0260) | 6.0555 | **<0.0001** | 0.0127 (0.0015 to 0.0239) | 2.2217 | **0.0263** |

<sup>a</sup>Multivariate meta-regression was performed when the univariate meta-regression P value was < 0.05.
<sup>b</sup>Duration from December 1, 2019 to the middle time of study period.
<sup>c</sup>Mean time from symptom onset to hospital admission.

**Table 3.**
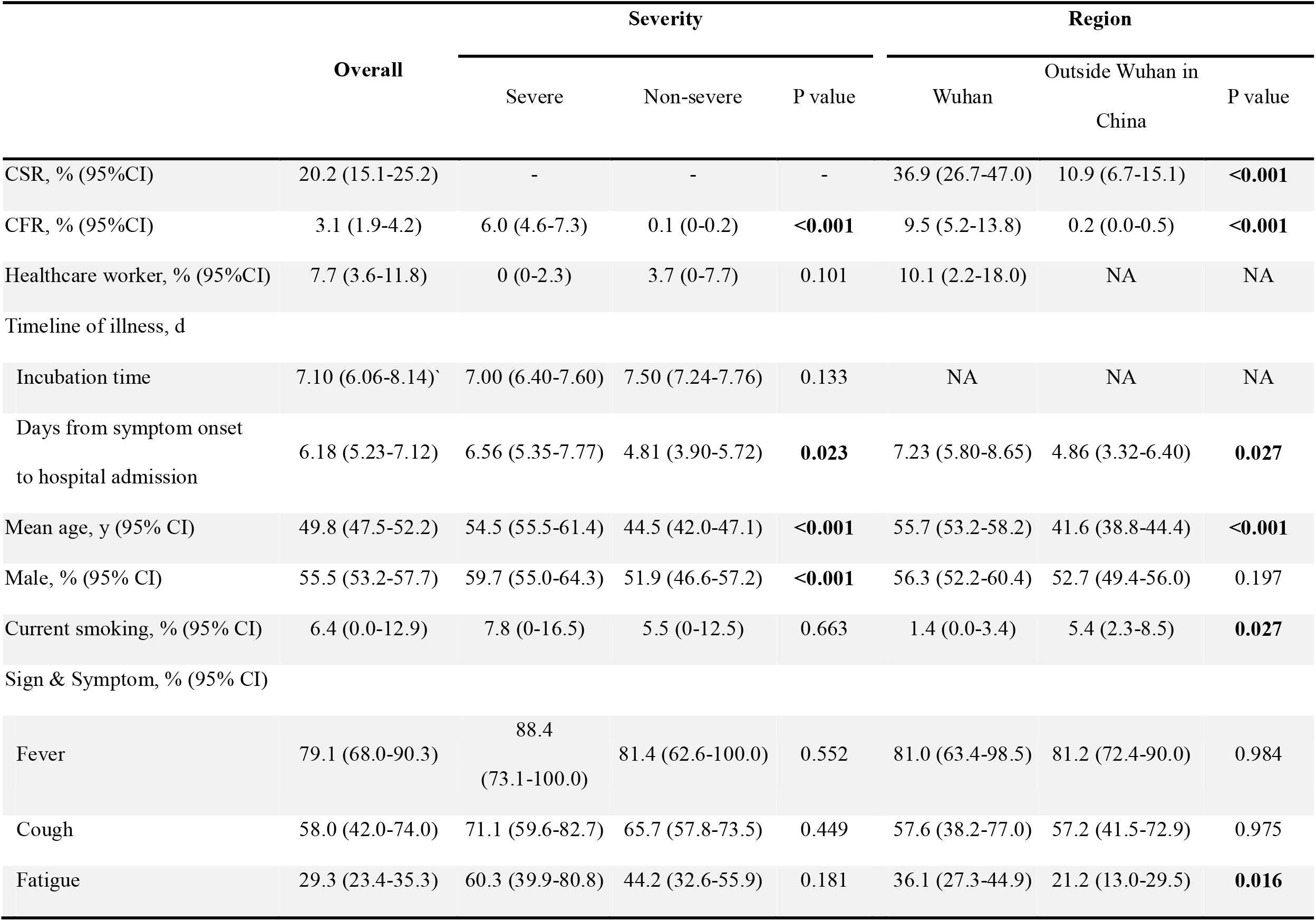
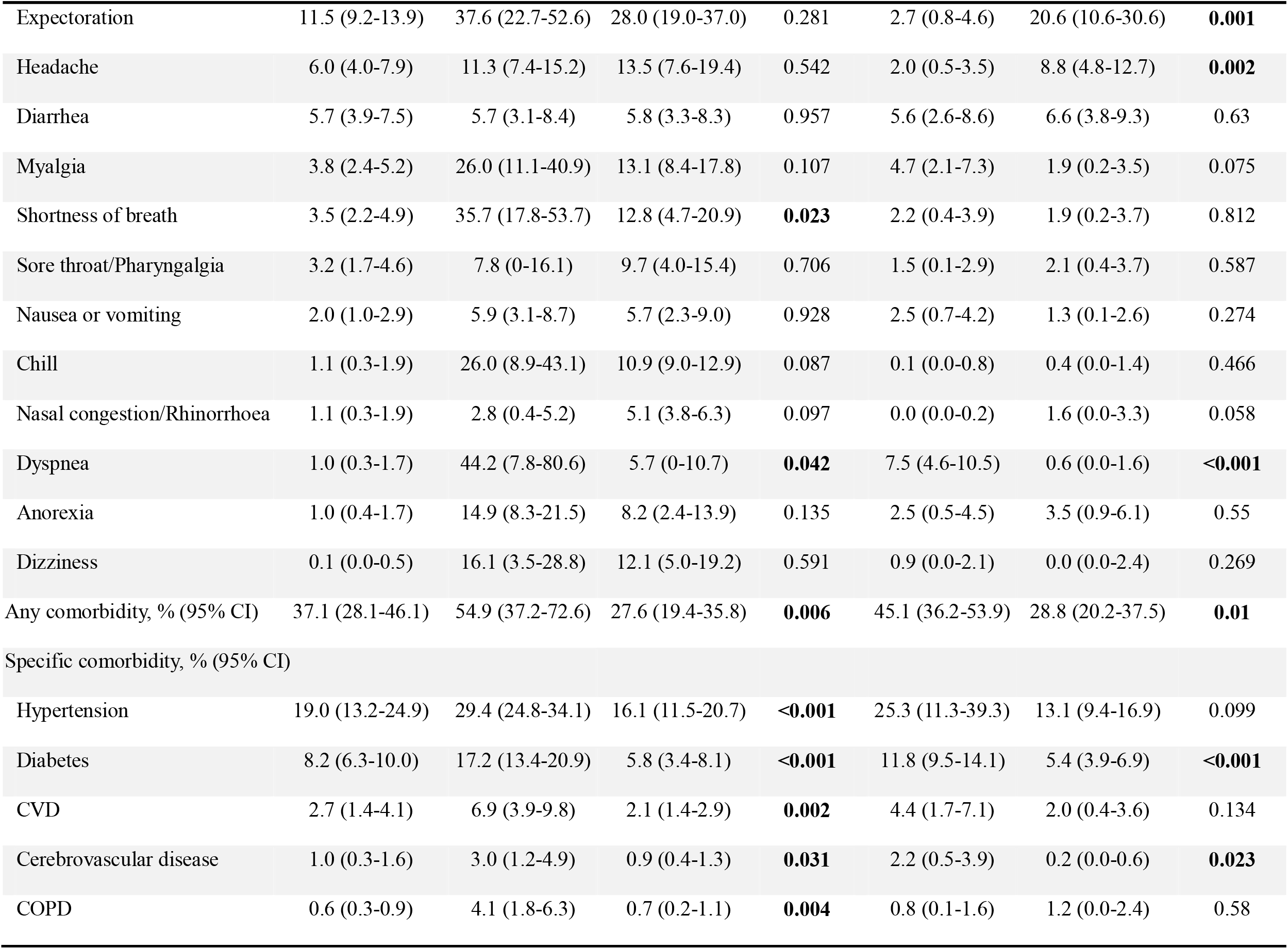
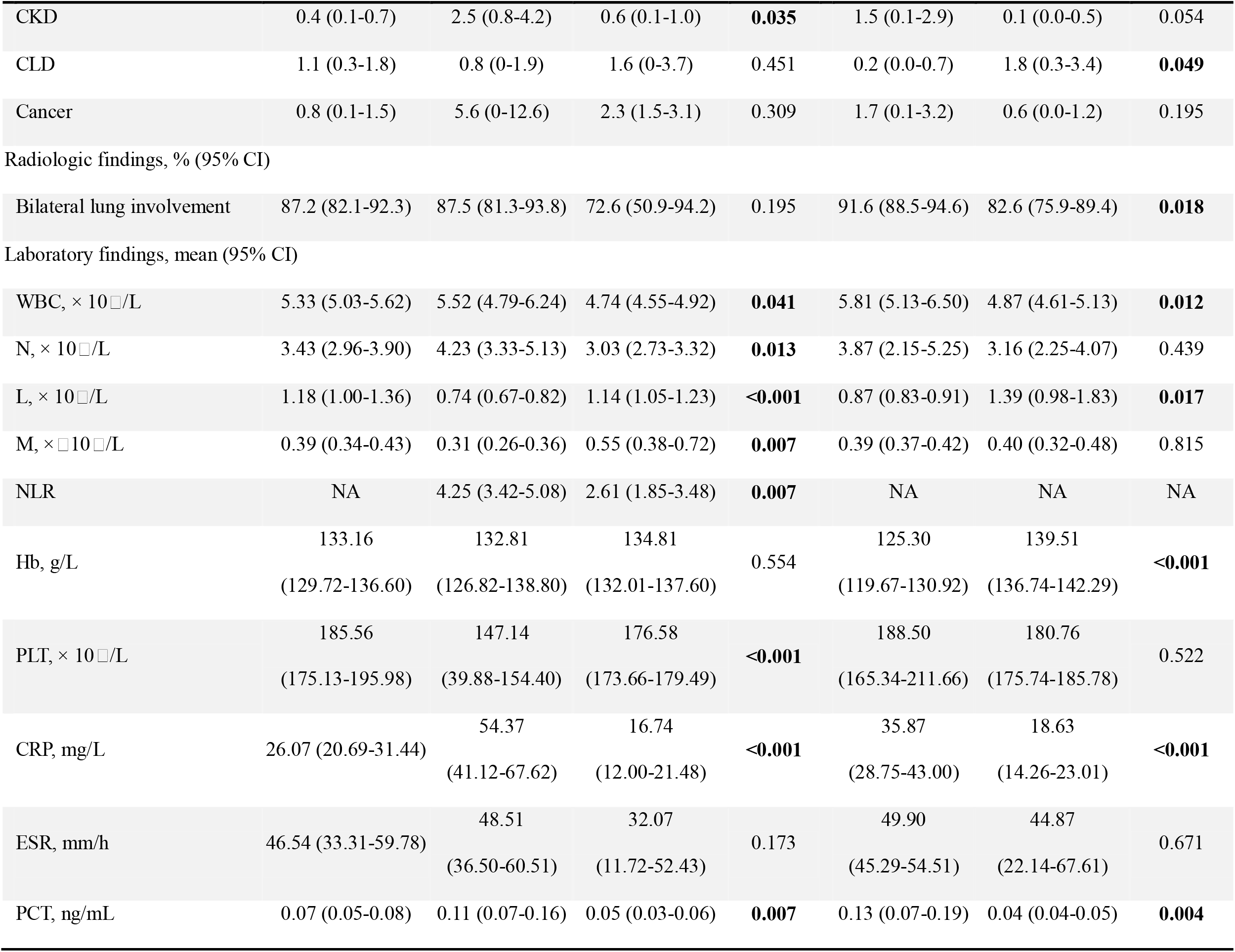
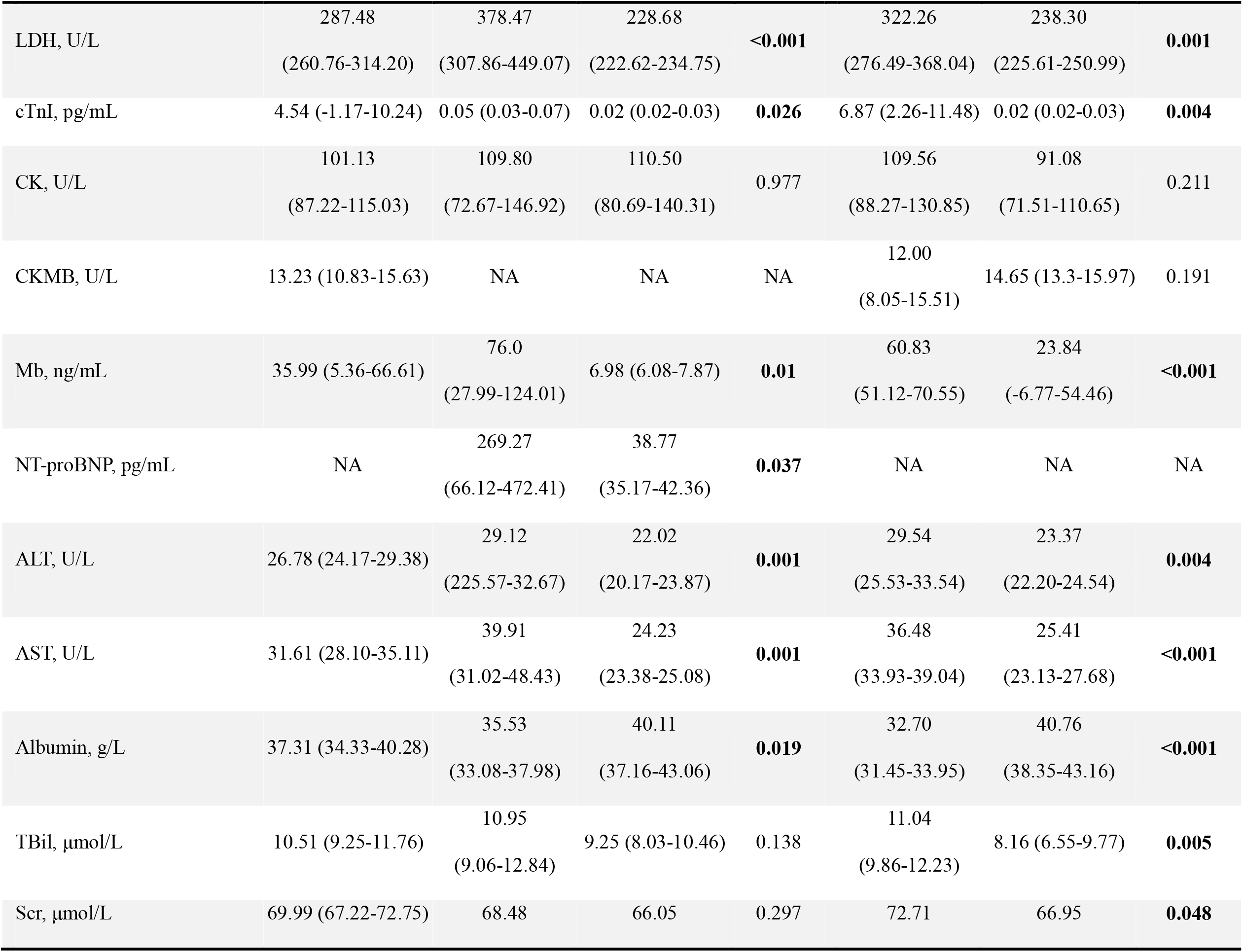
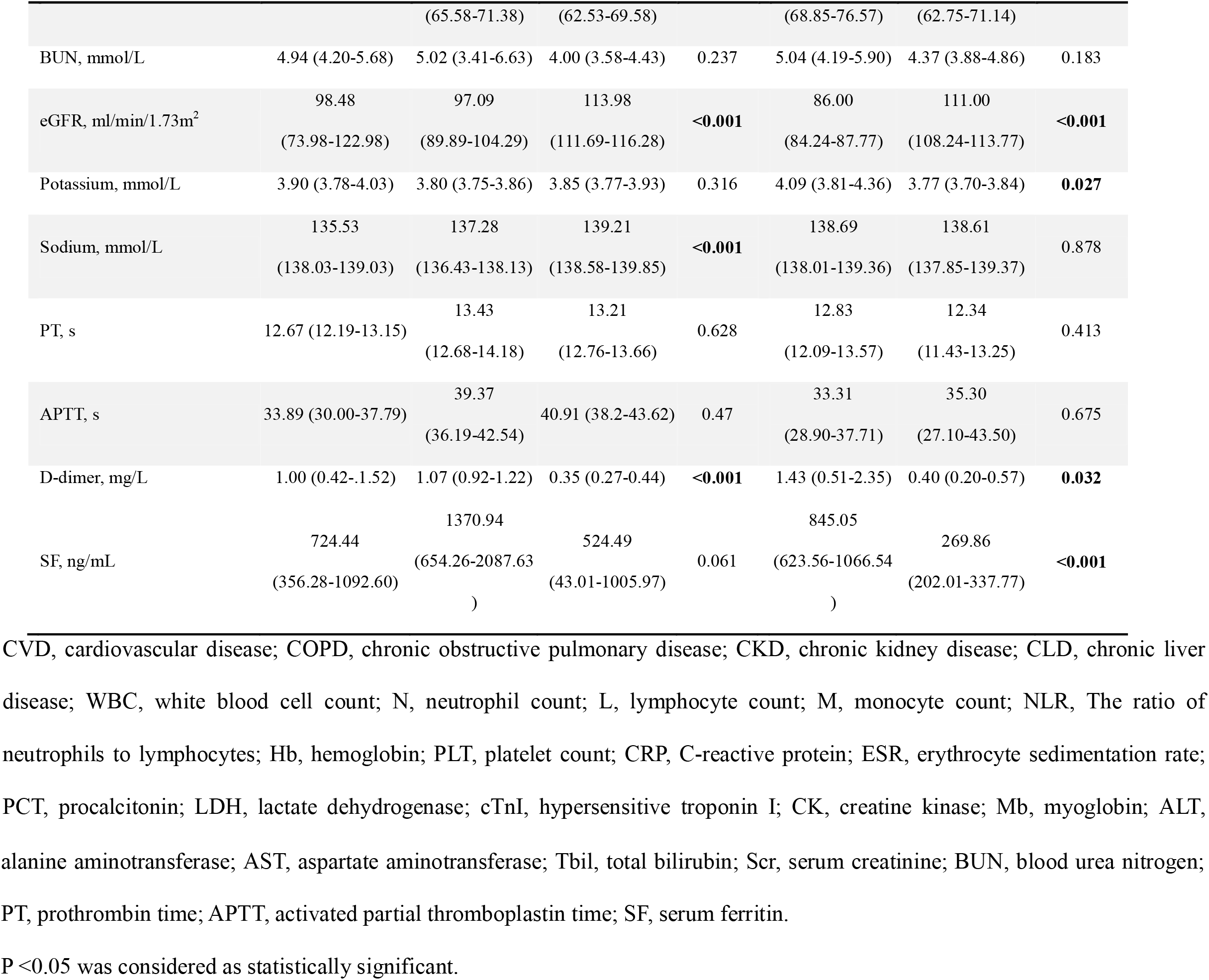
Clinical and laboratory data of the included COVID-19 patients.

**Figure 2.**
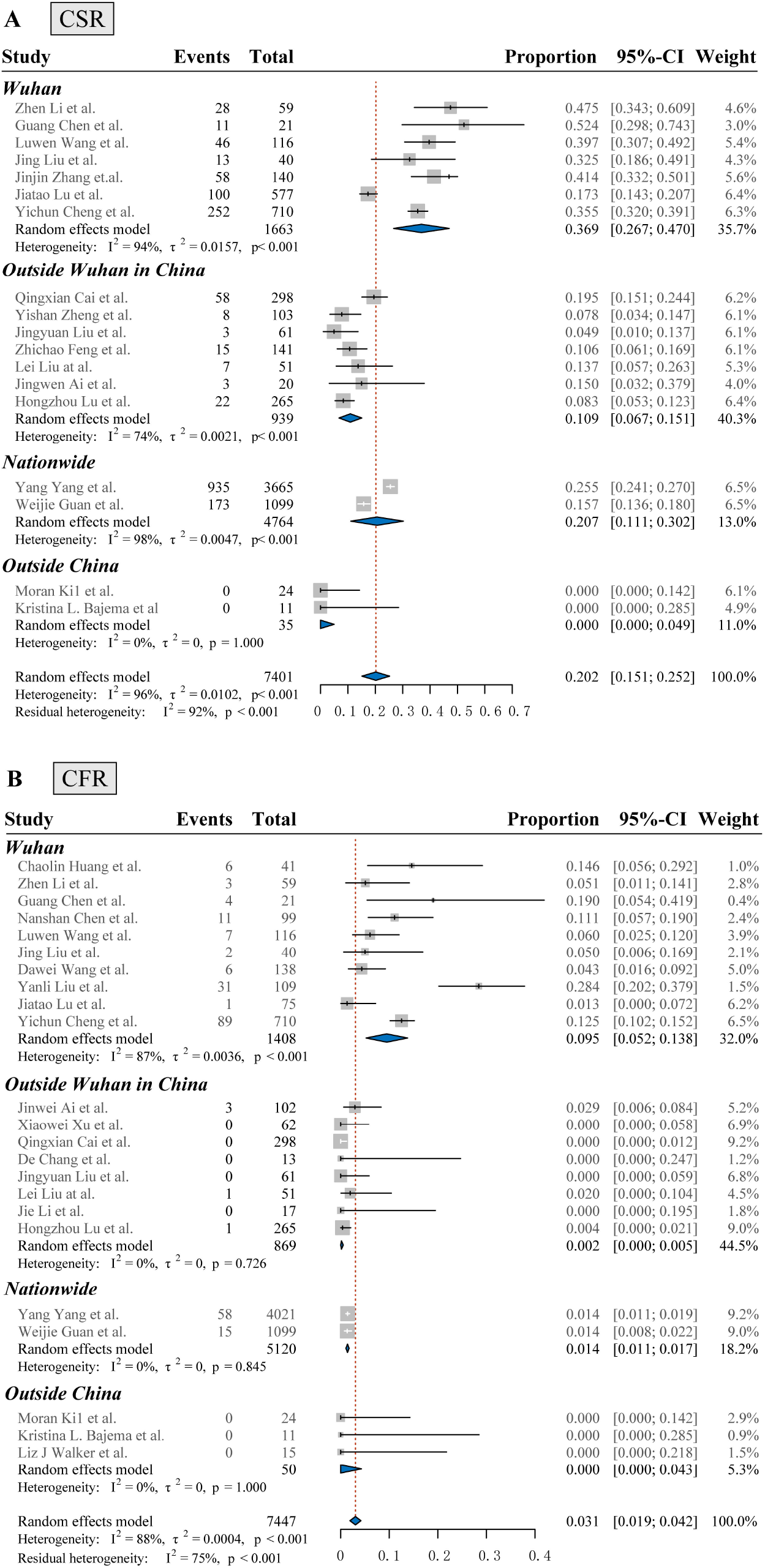
Subgroup analysis of CSR (A) and CFR (B) by geographical region. CSR, case severity rate; CFR, case fatality rate.

**Figure 3.**
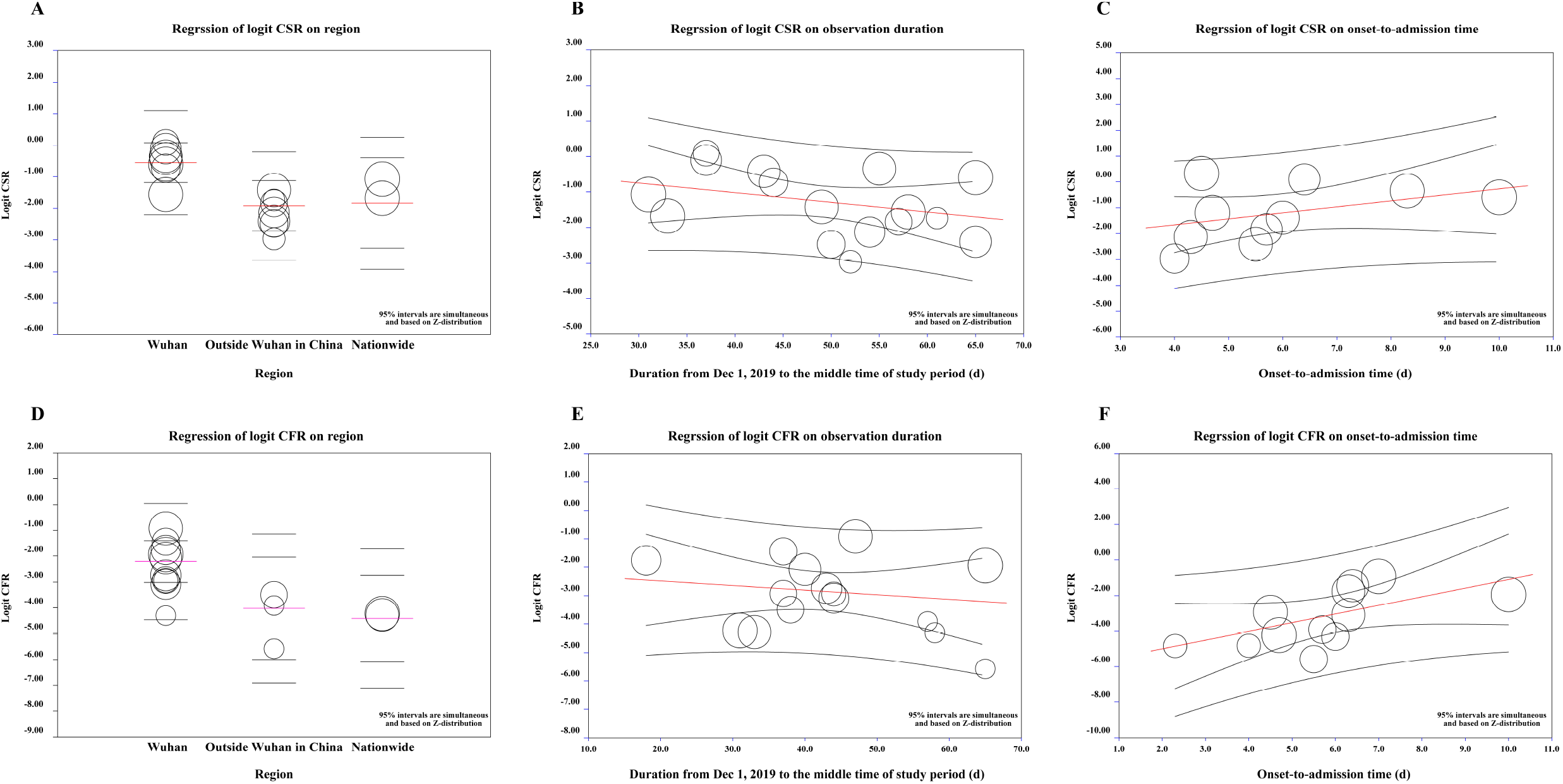
Meta-regression analysis for CSR and CFR. (A) meta-regression of Logit CSR on geographical region (Wuhan, outside Wuhan in China, nationwide). (B) meta-regression of Logit CSR on duration from Dec 1, 2019 to the middle time of study period. (C) meta-regression of Logit CSR on mean time from symptom onset to hospital admission (D) meta-regression of Logit CFR on geographical region (Wuhan, outside Wuhan in China, nationwide). (E) meta-regression of Logit CFR on duration from Dec 1, 2019 to the middle time of study period. (F) meta-regression of Logit CFR on mean time from symptom onset to hospital admission. The size of the circle represent the population size of each study. The red line in A&D represent the pooled incidence of CSR or CFR in each region, and the red line in B-C, E-F represent the regression equation from meta-analysis.

### Clinical characteristics and laboratory results

Clinical and laboratory data from 26 studies, including 1374 severe and 4326 non-severe patients, were extracted for meta-analysis. Of this, 7.7% patients (95% CI, 3.6-11.8%) were medical staff. The pooled CFR of severe patients was significantly higher than non-severe patients (6.0%; [95% CI, 4.6-7.3%] vs. 0.1%; [95% CI, 0-0.2%], *P* < 0.001). The mean incubation period was 7.10 days (95% CI, 6.06-8.14 d), with no statistically difference between severe and non-severe cases. The mean time from symptom onset to hospital admission was 6.18 days (95% CI, 5.23-7.12 d), which is longer in severe cases than that in non-severe cases (6.56 d vs. 4.81d, P = 0.023) and in Wuhan than outside (7.23 d vs.4.86 d, P = 0.027).

Severe cases presented with older age (54.5 yrs; [95% CI, 55.5-61.4 yrs] vs. 44.5 yrs; [95% CI, 42.0-47.1 yrs], *P* < 0.001) and higher ratio of male (59.7%; [95% CI, 55.0-64.3%] vs. 51.9; [95% CI, 46.6-57.2%], *P* < 0.001), as compared with non-severe cases. Similarly, patients in Wuhan were older than those outside Wuhan (55.7 yrs vs. 41.6 yrs, *P* < 0.001). Fever (79.1%; 95% CI, 68.0-90.3%), cough (58.0%; 95% CI, 42.0-74.0%) and fatigue (29.3%; 95% CI, 23.4-35.3%) were the dominant symptoms, whereas digestive symptoms such as diarrhea (5.7%; 95%CI 3.9-7.5%), nausea or vomiting (2.0%; 95% CI, 1.0-2.9%) were relatively rare. The overall prevalence of any comorbidity was 37.1% (95% CI, 28.1-46.1%), with higher incidence rate in severe cases than non-severe cases (54.9% vs. 27.6%, P = 0.006), and higher in Wuhan than other areas (45.1% vs. 28.8%, *P* = 0.01). The most common comorbidities were hypertension (19.0%, 95% CI, 13.2-24.9%), followed by diabetes (8.2%, 95% CI, 6.3-10.0%) and cardiovascular diseases (CVD, 2.7%, 95% CI, 1.4-4.1%). Moreover, hypertension, diabetes, CVD, cerebrovascular diseases, chronic obstructive pulmonary disease (COPD) and chronic kidney disease (CKD) were significantly more common in severe cases as compared with non-severe cases (all for *P* < 0.05). The overall proportion of bilateral radiologic abnormalities was 87.2% (95% CI, 82.1-92.3%), with significant difference between inside and outside Wuhan (91.6% vs. 82.6%, *P* = 0.018).

In terms of the laboratory index, several elevated indicators were observed as follows: C-reaction protein (CRP, 26.07 mg/L; 95% CI, 20.69-31.44 mg/L), erythrocyte sedimentation rate (ESR, 46.54 mm/h; 95% CI, 33.31-59.78 mm/h), lactate dehydrogenase (LDH, 287.48 U/L; 95% CI, 260.76-314.20 U/L), cardiac troponin I (cTnI, 4.54 pg/mL,; 95% CI, −1.17-10.24 pg/mL) and ferritin (724.44 ng/mL; 95%CI, 356.28-1092.60 ng/mL). In contrast, the level albumin (37.31 g/L; 95% CI, 34.33-40.28 g/L) and lymphocytes count (1.18 × 10□/L; 95% CI, 1.00-1.36 × 10□/L) were below normal level. Of note, obvious differences in laboratory index were identified between severe and non-severe cases, as well as between Wuhan and outside Wuhan. Elevated level of CRP, LDH and D-dimer, together with reduced level of lymphocytes count and PLT count were the prominent features of severe cases (all for *P* < 0.001). Likewise, more elevated CRP, myoglobin, aspartate aminotransferase (AST) and ferritin, followed by decreased lymphocyte count and hemoglobin were observed in Wuhan patients than outside (all for *P* < 0.001) (Table *2*).

### Risk factors for severity of COVID-19

Among the baseline characteristics, disease severity was highly associated with old age (≥ 50 yrs, OR = 2.609; 95% CI, 2.288-2.976; n = 5; I^2^ = 37%), male (OR =1.348; 95% CI, 1.195-1.521; n = 13; I^2^ = 0%), smoking (OR =1.734; 95% CI, 1.146-2.626; n = 4; I^2^ = 0%) and any comorbidity (OR = 2.635; 95% CI, 2.098-3.309; n = 7; I^2^ = 12%). Comorbidities with pooled OR larger than 2 included CKD (6.02; 95% CI, 2.19-16.51; n = 4; I^2^ = 0), COPD (5.32; 95% CI, 2.61-10.85; n = 6; I^2^ = 0%), cerebrovascular diseases (3.19; 95% CI, 1.51-6.77; n = 6; I^2^ = 0%), tumor (3.21; 95% CI, 1.42-7.24; n = 4; I^2^ = 30%), diabetes (2.49; 95% CI, 1.82-3.4; n = 10; I^2^ = 44%) and hypertension (2.06; 95% CI, 1.61-2.62; n = 10; I^2^ = 36%) (Figure *4*, eFigure *5*).

**Figure 4.**
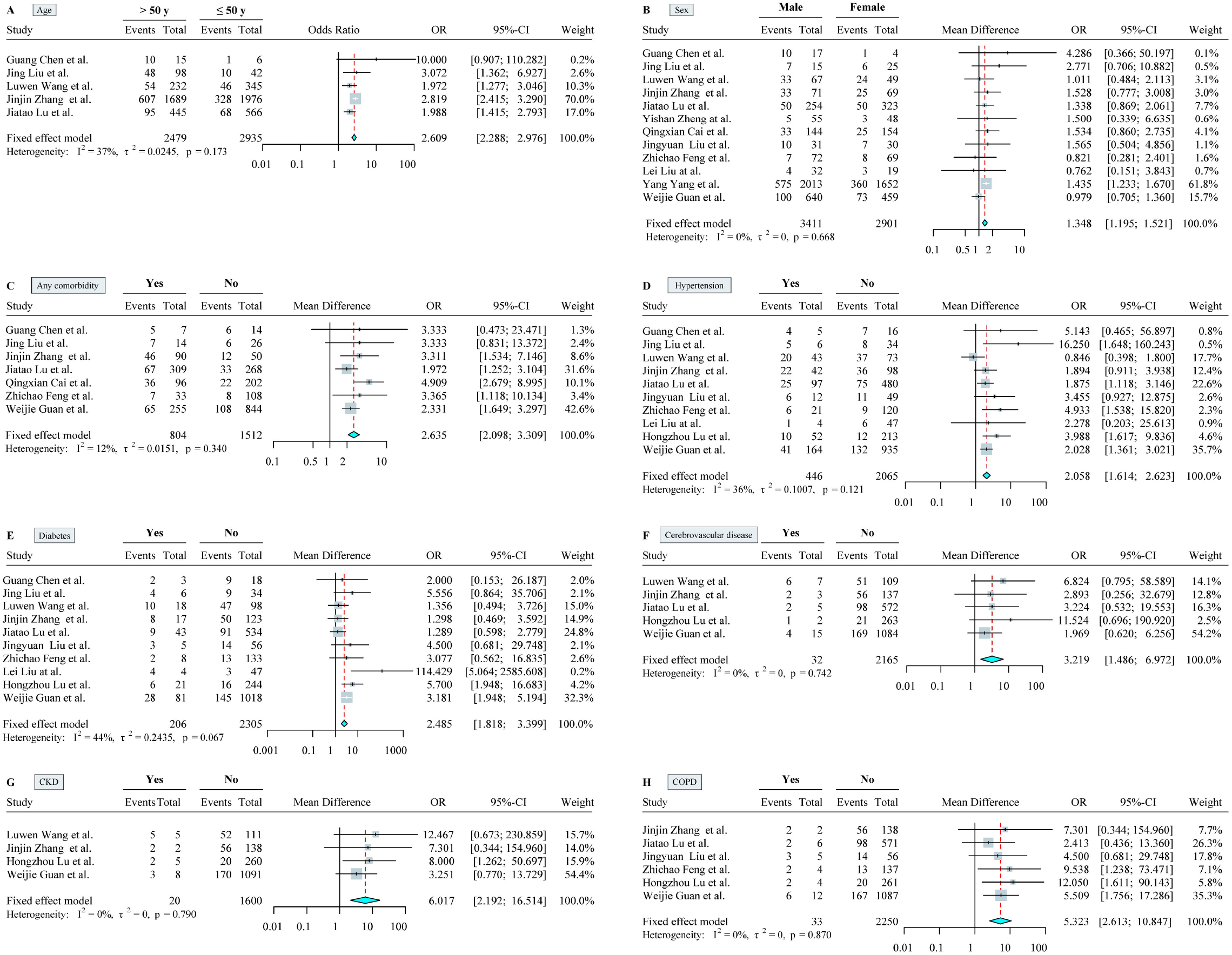
Clinical risk factors for severe cases. A, old age (> 50 yrs); B, male gender; C, any comorbidity; D, hypertension; E, diabetes; F, cerebrovascular diseases; G, chronic kidney diseases (CKD); H, chronic obstructive pulmonary diseases (COPD).All factors were dealt as dichotomous variables and odds ratios (ORs) were showed using fixed effect model (all for I^2^ < 50%).

In terms of laboratory results, there were obvious difference between severe and non-severe cases in PLT (MD = −30.654 × 10^9^/L, 95% CI, −38.7 to −22.61, n = 8), lymphocyte count (MD = −0.376 × 10^9^/L, 95% CI, −0.467 to −0.285, n = 11), LDH (MD = 150.702 U/L, 95% CI, 82.569 to 218.836, n = 5), D-dimer (MD = 0.715 mg/L, 95% CI, 0.562 to 0.868, n = 7) and CRP (MD = 30.395 mg/L, 95% CI, 20.006 to 40.784, n = 10). Meanwhile, the ORs were calculated for several index including lymphocytopenia (OR = 4.23; 95% CI, 3.03-6.03; I^2^ = 33%), thrombocytopenia (OR = 2.84; 95% CI, 2.00-4.04; I^2^ = 49%), elevated D-dimer (OR = 3.17; 95% CI, 1.86-5.41; I^2^ = 69%) and elevated CRP (OR = 4.23; 95% CI, 2.94-6.08; I^2^ = 25%). (Figure *5*, eFigure *6-7*).

**Figure 5.**
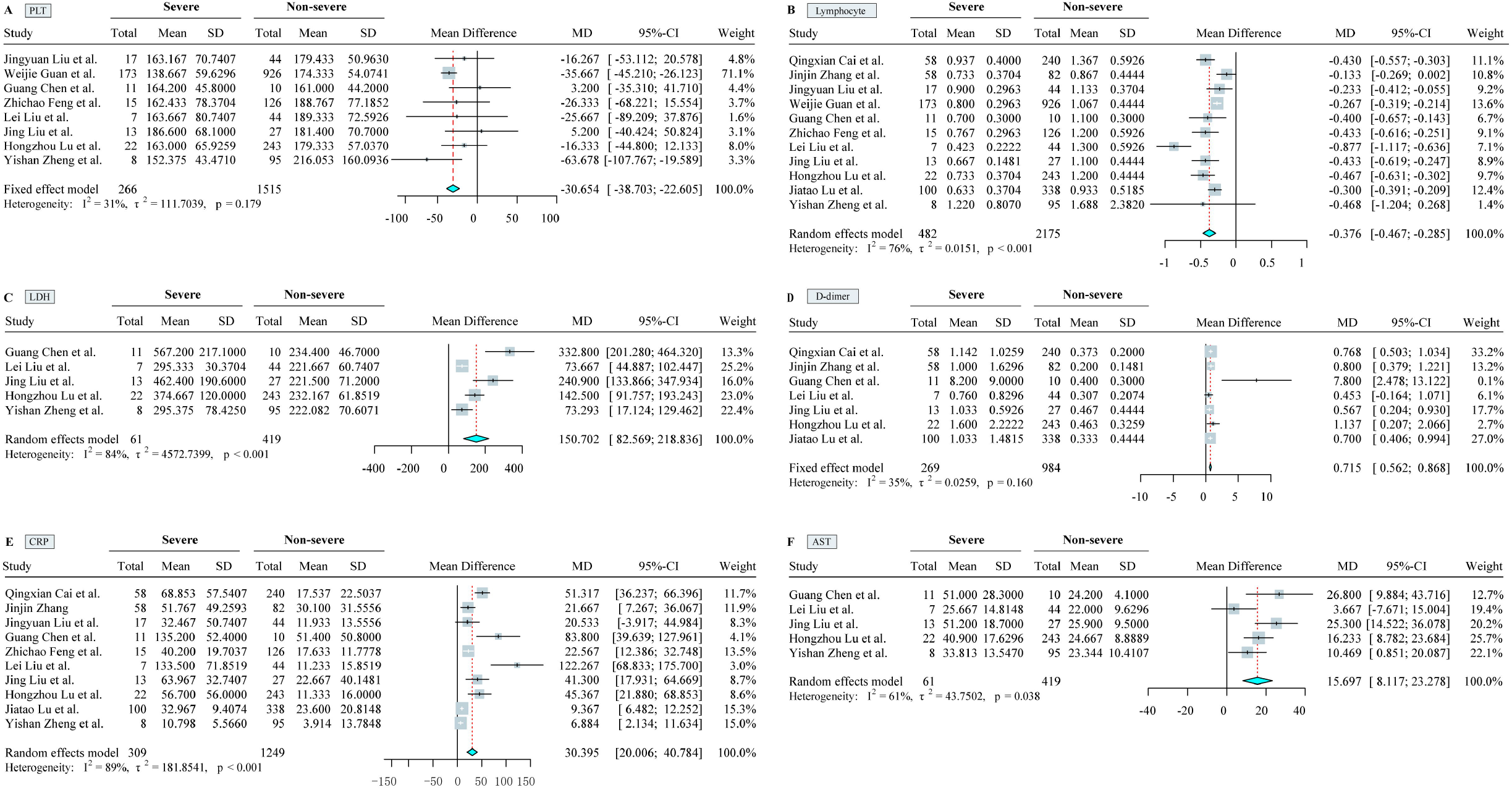
Mean Difference (MD) in main laboratory index between severe and non-severe patients with COVID-19. A, blood platelet count (PLT, × 10^9^/L); B, lymphocyte count (× 10^9^/L); C, lactate dehydrogenase (LDH, U/L); D, D-dimer (mg/L); E, C-reaction protein (mg/L); F, aspartate aminotransferase (AST, U/L).

### Comparison of Clinical Characteristics and risk factors among COVID-19, SARS and MERS

The most common comorbidity was diabetes for both SARS (24.0%) and MERS (68.0%), while hypertension for COVID-19 (19.0%). Fever and cough were the dominant symptoms for all three viruses, whereas digestive symptoms and chill were relatively rare for COVID-19. Regarding laboratory index, elevated LDH was common for three coronaviruses; higher incidence of lymphopenia, elevated AST or ALT was observed in COVID-19 and SARS than MERS^39,40^. (Table 4). Both old age and comorbidity proved the common risk factors for predicting death among three coronaviruses. In support, COVID-19 related death was associated with old age (≥ 60 yrs, RR = 9.45; 95% CI, 8.09-11.04), male (RR = 1.67, 95% CI, 1.47-1.89) and any comorbidity (5.86; 95% CI, 4.77-7.19), most notably CVD (6.75; 95% CI, 5.40-8.43) followed by hypertension (4.48; 95% CI, 3.69-5.45) and diabetes (4.43; 95% CI, 3.49-5.61). Similar with COVID-19, male (RR = 1.6; 95% CI, 1.2-2.1) and CVD (OR = 3.5; 95% CI, 3.1-4.8) were also risk factors for MERS related death^41,42^. In contrast, the most predominant risk factor for SARS related death was CKD (9.02; 95% CI, 3.81-21.36), and the risk of male gender was not statistically significant^3,43,44^. In addition, medical staff had a lower fatality rate than non-clinical staff for COVID-19 (RR = 0.12; 95% CI, 0.05-0.30) and MERS (RR = 0.1; 95% CI, 0.02-0.20), whereas the difference was not significant for SARS (RR = 0.76; 95% CI, 0.52-1.15). (Table *5*).

**Table 4.** Comparison of clinical characteristics among COVID-19, SARS and MERS.

| Clinical Characteristics | COVID-19 | SARS | MERS |
| --- | --- | --- | --- |
| CFR, % | 3.1 | 9.6 | 40 |
| Mean age, y | 49.8 | 39.9 | 50 |
| Male, % | 55.5 | 43 | 64.5 |
| Health care worker, % | 7.7 | 23 | 9.8 |
| Current smoking, % | 6.4 | 17 | 23 |
| Any comorbidity, % | 37.1 | 10-30 | 76 |
| Diabetes | 8.2 | 24 | 68 |
| Chronic renal disease | 0.4 | 2-6 | 49 |
| Chronic heart disease | 2.7 | 10 | 28 |
| Cancer | 0.8 | 3 | 2 |
| Hypertension | 19 | 19 | 34 |
| Presenting symptoms, % |  |  |  |
| Fever | 79.1 | 99-100 | 98 |
| Cough | 58 | 62-100 | 83 |
| Diarrhea | 5.7 | 20-25 | 26 |
| Myalgia | 3.8 | 45-61 | 32 |
| Shortness of breath | 3.5 | 40-42 | 72 |
| Chill | 1.1 | 15-73 | 87 |
| Laboratory results, % |  |  |  |
| Leucopenia ($<4.0 \times 10^9$ /L) | 22.2 | 25-35 | 14 |
| Lymphopenia ($<1.5 \times 10^9$ /L) | 54.7 | 68-85 | 32 |
| Thrombocytopenia ($<140 \times 10^9$ /L) | 12.8 | 40-45 | 36 |
| High LDH ($>245$ U/L) | 42.9 | 50-71 | 48 |
| High ALT (>40 U/L) | 20 | 20-30 | 11 |
| High AST (>40 U/L) | 23.8 | 20-30 | 14 |

**Table 5.**
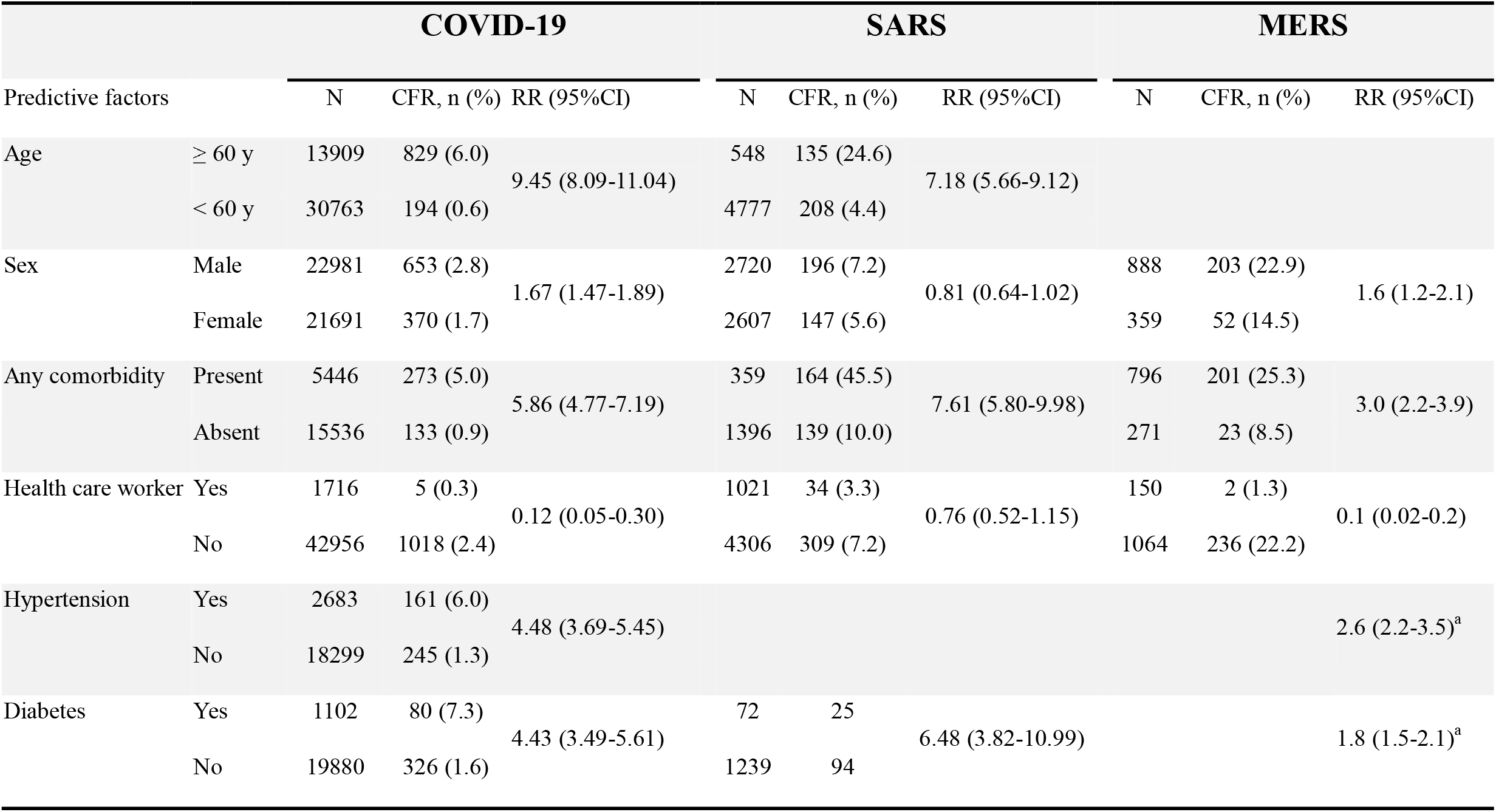
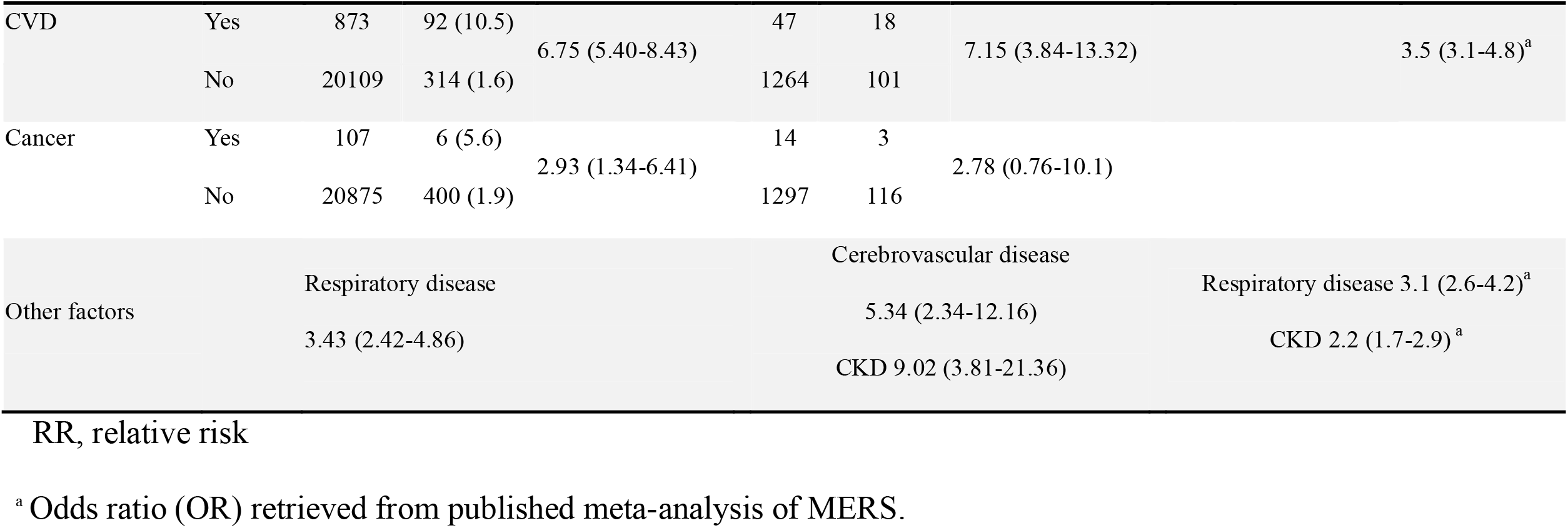
Comparison of risk factors for death among COVID-19, SARS and MERS.

### Sensitivity Analysis and Publication Bias

Leaving each trial out of the analysis one at a time revealed no meaningful differences in CSR and CFR (eFigure *8* in supplement). We observed no evidence of publication bias with inspection of the funnel plot or with the Begger test or the test used by Peters et al (eFigure *9* in supplement).

## Discussion

The main findings of this present analysis are that: (1) Despite the high incidence rate, the distinctive feature of COVID-19 was low severity and mortality, showing a significant difference on the incidence of severity and mortality between Wuhan and outside of Wuhan; (2) The onset-to-admission time was closely related to mortality, which will be increased about 1.27% with every day of delay in admission; (3) Older age is a common risk factor for poor prognosis of the three coronavirus diseases; Hypertension and cardiovascular diseases are the most relevant clinical predictors for the death caused by both COVID-19 and MERS-CoV, while chronic kidney disease is for SARS; (4) The five main laboratory indicators of severe illness included lymphocytopenia, thrombocytopenia and the rise of LDH, CRP and D-dimer.

SARS-CoV-2 has been reported to be higher contagious than previously discovered human coronaviruses^45^. Until now, more than 187, 361 confirmed cases and caused 7, 485 deaths in 151 countries on six continents were identified. Despite its high prevalence of COVID-19, the pooled severe incidence and fatality rate is significantly lower compared with SARS and MERS, which may explain why the novel coronavirus has spread so widely^46^. Of note, there are regional and spatial differences in the incidence rate of COVID-19. In our research, the pooled severity rate and mortality caused by COVID-19 was found significantly higher in Wuhan than that of the infected outside of Wuhan (all for *P* < 0.01). On the other hand, disease incidence at the early stage of outbreak was higher than that at the late stage, which may be caused by the lack of recognitions and treatment experience for COVID-19. Moreover, the longer time from symptoms to hospitalization, the higher incidence rate of the mortality related to COVID-19, highlighting the importance of timely medical treatment^30^. In addition, among the patients with 2019-nCoV, the pooled infection rate of medical staff was 7.7%, which was lower than that of SARS (23%) and MERS (9.8%)^39^.

SARS-CoV-2, SARS-CoV and MERS-CoV belong to high pathogenic coronaviruses, whereas each of which has its own clinical manifestation. In comparison to SARS-CoV (4.6 days) and MERS-CoV (5.2 days)^47^, COVID-19 has a longer latent period (7.1 days) and the initial manifestations are non-specific, making it more difficult to prevent and control at early stage. Similar to SARS and MERS, persons with COVID-19 often present initially with lower respiratory signs, including fever, coughing and fatigue. In the late course of illnesses, infected persons are characterized by progressive breathing difficulty, tachypnea, acute respiratory distress syndrome, or life-threatening complications. In fact, SARS-CoV-2 has been isolated from respiratory secretions, feces, urine, blood, tears, and conjunctival secretions^48^, indicating that SARS-CoV-2 infection is not confined to the respiratory tract. Indeed, in our analysis, the pooled incidences rate of respiratory symptom was present in 79.1% of patients, followed by 7.7% with gastrointestinal disorders and 6.1% with neurological symptoms. Therefore, the patients with COVID-19 whose initial symptom were out of the lung should also be paid more attention, especially for those with the contact history of COVID-19. It should be noted that the pooled incidence of diarrhea symptoms (5.7%) is lower than previous data of patients with MERS-CoV (25%) or SARS-CoV (26%) infection. In terms of laboratory tests, the most common hematological abnormalities in patients with COVID-19 were lymphopenia (54.7%), suggesting aggressive effect on lymphocytes by COVID-19, which is similar to those previously observed in patients with SARS (68-85%). In addition, elevated levels of liver enzymes, LDH, myocardial enzymes, and depressed platelet count concomitant with the rise of D-dimer was observed in our study. Compare with MERS (36%) and SARS (45%), thrombocytopenia was relatively less frequent in patients infected with SARS-CoV-2 (12.8%)^39,47^.

Given the fact that the rapid progression to end-organ failure and even death was occurred in some patients, it is therefore essential to paid more attention to susceptible population of COVID-19^49^. Our results showed the majority of the SARS-CoV-2 infection were male patients (55.5%) similarly to the gender distribution of MERS (64.5%), while the predominance of female patients (43.0%) was observed in SARS. It is believed that the sex difference is probably related to the higher expression of ACE2 receptor in male than that in female and the lack of the protection of estrogen and X chromosome^50^. COVID-19 has affected persons in all age groups; in particular, 53.5% patients were found after more than 50 years, suggesting that the elderly patients are more likely to have weak immune function. Moreover, 10-30% of patients in SARS and 37.1% of patients in COVID-19 had at least one underlying disorder; patients with comorbidities such as diabetes mellitus, cardiovascular diseases, renal failure and chronic respiratory diseases are especially vulnerable to SARS-CoV-2 infection^7^. Owning to the pro-inflammatory state and the reduced immune response, the chronic conditions were also noted to have similar effects in the other two coronaviruses. Although the comorbidities identified in our study has been described previously, their value to predict the severity of COVID-19 has not yet been evaluated.

A focus on risk factor affecting clinical outcome is critically important to identify high-risk patients and mitigate COVID-19 complications. In the present study, people with old age, male, smoking, presence of comorbidity, CKD, COPD, cancer, hypertension, and diabetes were identified as predictors of severe disease from COVID-19 infection. Importantly, old age, male gender and presence of comorbidity, including hypertension, diabetes, cancer and respiratory disease, were identified predictor of disease severity as well as mortality related to COVID-19, suggesting that elderly patients with these underlying comorbidities should be given more attention and care. In line with previously published studies, the laboratory indicators including lymphocytes, CRP, LDL, PLT, D-dimer, ALT and CK levels were closely related to a poor prognosis, which provided key reference index of the prognosis of COVID-19. In particular, lymphopenia, thrombocytopenia and elevated D-dimer could act as effective predictors of the COVID-19 severity. However, it was worth noting that increased level of myoglobin and ferritin were observed in severe cases, but the predictive value could not be estimated limited by the number of related studies and further in-depth research is needed.

## Limitation

Our meta-analysis has several potential limitations. Firstly, there was obvious heterogeneity among studies regarding CSR and its subgroups, because of differences in medical condition, study period, public awareness and others. Nevertheless, we conducted meta-regression based on the observation duration and symptom onset to hospital admission time, which explained a large percent of heterogeneity. Secondly, studies published before February 25, 2020 and articles published in English only were included in our study, therefore there was lack of data from other countries. However, our meta-analysis involved 53000 confirmed patients based on the data during the early-to-mid period of disease outbreak in China, which will provide great referential value for global epidemic control. Thirdly, meta-analysis was conducted on the level of the studies and the characteristics of individual patients could not be retrieved, thus it was hard to provide reference for individualized diagnosis and treatment of COVID-19. Finally, all included studies were retrospective, as no randomized control trials and prospective studies related to 2019-nCoV finish till now, thus our results require to be confirmed by more high-quality clinical researches.

## Conclusions

COVID-19 is emerging all over the world and spreading at an unprecedented rate, resulting in significant impacts on global economies and public health. The present study successfully and systematically evaluated the prognostic predictors of COVID-19 by collecting published information on risk factors of the outcomes related to SARS-CoV-2 infections. Nevertheless, more investigations are needed to further objectively confirm the clinical value of prognostic factors related to COVID-19.

## Data Availability

The raw/processed data required to reproduce these findings cannot be shared at this time as the data also forms part of an ongoing study.

